# VERITAS: A Neuro-Symbolic Approach to Quantifying Epistemic Divergence and Harm Potential in Online Health Narratives

**DOI:** 10.64898/2026.07.11.26357840

**Authors:** Ommo Clark, Anupam Joshi, Karuna Pande Joshi

## Abstract

**Objective:** Online health information seeking is rising, and individuals increasingly act on peer advice without clinical oversight, adjusting doses, delaying care, and modifying treatment. Current misinformation detection assumes factually inaccurate content is what makes these decisions unsafe. We introduce VERITAS (**V**erification **E**ngine for **R**isk-aware **I**nformation **T**rust **A**ssessment in health **S**tories) and formalize the Risk Irrelevance Principle: divergence from accepted clinical practice and potential for harm are distinct, weakly associated dimensions that must be assessed separately.

**Materials and Methods:** VERITAS transforms unstructured health narratives into Agent-Action-Outcome graphs and computes two continuous metrics: Narrative Truth Distance (NTD), quantifying epistemic divergence, and Narrative Risk Score (NRS), assessing harm potential. We evaluated VERITAS on 704 threads from four Reddit health communities. Two domain experts annotated 2,000 segments (Krippendorff’s α=0.78-0.81). NTD-NRS independence was validated using seven tests.

**Results:** NTD and NRS shared under 5% of variance (r = 0.222; mutual information 0.096 bits): a post’s divergence from consensus conveys little about whether acting on it will cause harm. On 435 labeled posts, VERITAS identified 62.2% of expert-labeled misinformation versus 57.5% for the strongest text classifier, the gain concentrated in factually plausible content describing unsafe self-management (27.6% of misinformation) that accuracy-focused classifiers approve. VERITAS assessed 37.8% of this misinformation as low-risk, pending clinical validation.

**Discussion:** Fact-checking-based screening systematically approves the content most likely to prompt unsafe self-management while flagging content least likely to cause harm.

**Conclusion:** Separating divergence from harm potential shifts verification from whether information is correct to whether it is safe to act upon.

## INTRODUCTION

### Background And Significance

Online platforms have become primary sources of health information, with individuals routinely relying on peer-generated narratives to guide self-care decisions outside formal clinical supervision.^1–3,9,23^ Our prior survey across seven countries found that 87% of participants expressed concern about health misinformation yet continued engaging in online health-seeking behaviors due to convenience, relatability, and barriers to healthcare access, with diabetes, hypertension, and weight management - conditions that incidentally require ongoing self-management between clinical encounters, the top three searched for.^4,5^

*We define online health safety as the protection of individuals from harm arising from health-related decisions in non-clinical settings guided by online information, where consequential decisions, medication adjustment, treatment modification, delayed care-seeking, occur without clinical supervision*. Existing frameworks define health misinformation around accuracy rather than safety,^61,62^ and traditional patient safety assumes clinician mediation within formal healthcare systems^58–60^; online health safety addresses the distinct reality that individuals act on peer-generated information without clinical oversight. These decisions directly influence treatment trajectories and health outcomes at both individual and population levels, creating a critical challenge for digital health systems and public health surveillance.^42^ Computational approaches to assessing online health information therefore represent critical public health infrastructure requiring the same rigor applied to clinical decision support systems.

However, user-generated health information online is shared as narratives, not isolated facts. People describe who did what and what resulted, stories with agents, actions, outcomes, and causal reasoning. Current computational systems exhibit what we have termed narrative blindness; the inability to read the causal structure and contextual inferences determining whether information is safe to act upon.^6–8,10^ This limitation is compounded by the detection paradigm that treats deviation from medical consensus as a proxy for harm^55–57^. Yet in non-clinical settings, health risk arises from how a narrative reasons and what it invites the reader to do: the causal claims it makes (for example, attributing symptom relief to stopping a medication), whether advice sound in one clinical context is transferred to another where it is unsafe, and how confidently claims are asserted relative to their supporting evidence. Accuracy-focused systems consequently miss content with correct medical facts describing unsafe actions while over-flagging harmless alternative practices.^6–8^ This risk-divergence misalignment, where content most likely to cause harm is least likely to be flagged, is particularly consequential in chronic disease contexts where online narratives frequently influence dosing, timing, and treatment modification.^4,5,34^

We term this the Risk Irrelevance Principle, that is, divergence from accepted clinical practice and harm potential are practically independent, by which we mean the association is too weak for either dimension to serve as a useful proxy for the other: the observed correlation (r = 0.222) falls below the |r| < 0.30 threshold pre-specified in our analysis plan, corresponding to Cohen’s convention for less than a medium effect, and to under 5% shared variance. A narrative recommending unsupervised insulin dose escalation using correct pharmacological terminology poses acute and/or severe health risk despite perfect epistemic alignment; conversely, a narrative describing a culturally traditional remedy using non-standard terminology may diverge substantially from established consensus while posing negligible health risk. Online health safety therefore requires risk-aware credibility assessment rather than truth-conditional classification.

## OBJECTIVE

This study has two aims: (1) to formalize and empirically validate the Risk Irrelevance Principle, and (2) to demonstrate that separating epistemic divergence from harm potential improves detection of safety-critical online health content. Building on our prior work formalizing narrative blindness and developing Agent-Action-Outcome (AAO) narrative graph,^10^ we introduce VERITAS (**V**erification **E**ngine for **R**isk-aware **I**nformation **T**rust **A**ssessment in health **S**tories), a framework that extends the concept of epistemic divergence^29,37^ to computational health informatics.

The Risk Irrelevance Principle is formalized as approximate statistical independence between epistemic divergence and potential harm, operationalized through mutual information and grounded in Shannon’s information theory^26,30,39^ and Halpern’s formalization of reasoning under uncertainty.^40^ Empirically, the low mutual information between the two dimensions (I(D;R) = 0.096 bits) indicates that a narrative’s epistemic divergence conveys little about its potential for harm.

For NTD, Shogenji’s formalization of epistemic justification^38^ motivates KL divergence as the operationalization of truth-distance: true belief systems exhibit decreasing divergence from ground truth as evidence accumulates, while internally coherent but false systems do not. Lycett and Partridge’s ontology-epistemology distinction^36^ separates shared medical vocabulary (factual divergence) from divergent causal reasoning (causal divergence), explaining why laypeople may use correct terminology yet reach harmful inferences, a pattern that confounds classifiers optimized for lexical verification. Confidence calibration theory^27^ grounds evidence divergence, the misalignment between assertion confidence and supporting evidence.

## MATERIALS AND METHODS

### Study Design and Reporting

VERITAS has a three-phase architecture: (1) narrative graph construction, (2) three-pillar verification engine, and (3) two-dimensional NTD and NRS scoring metrics and classification. We evaluated VERITAS as a risk-aware classification system for online health narratives and compared its performance with binary NLP misinformation approaches. Study design, validation, and reporting followed TRIPOD guidelines.^33^

**Figure 1.**
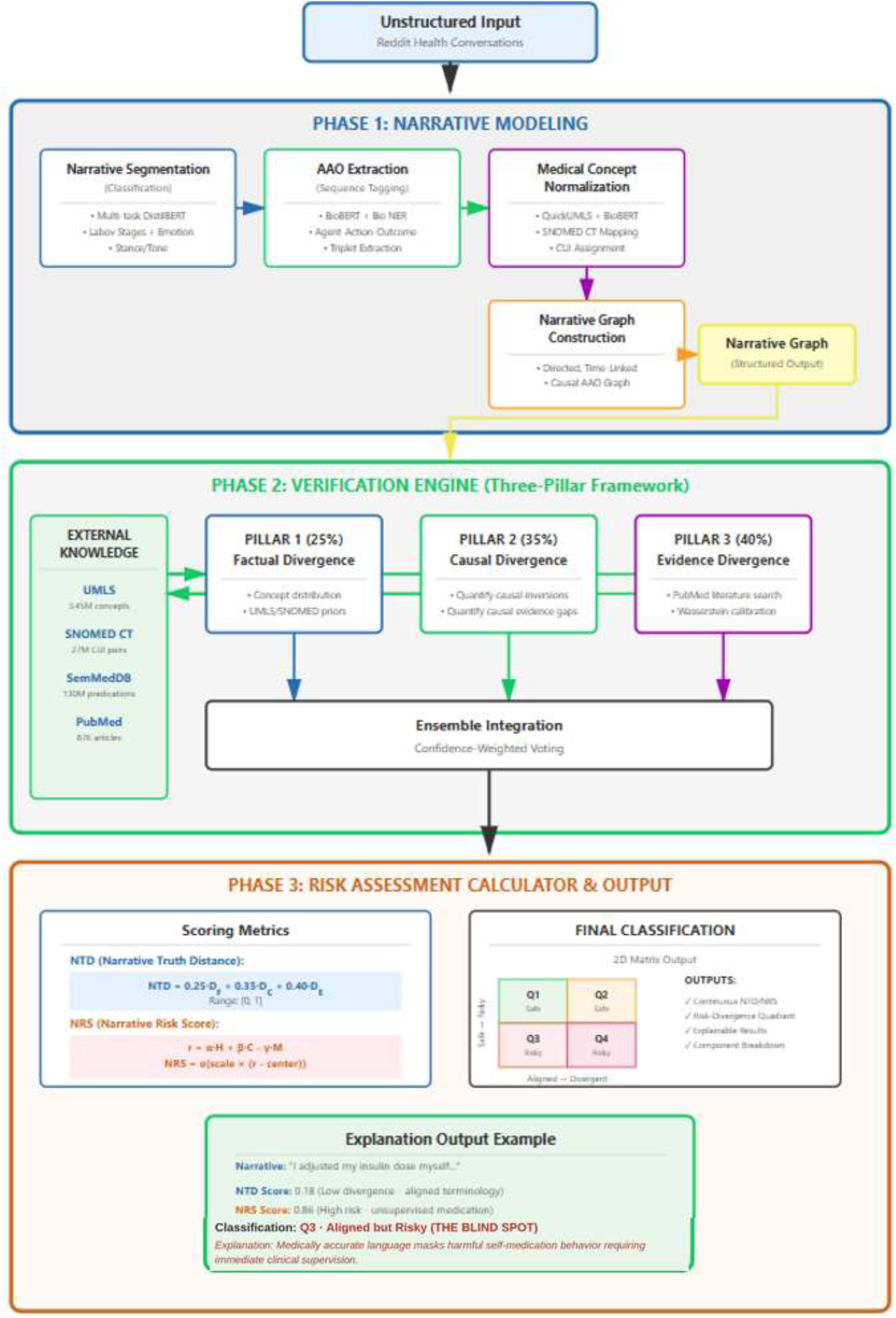
VERITAS Three-phase Pipeline. Flowchart of the VERITAS three-phase pipeline: Phase 1 constructs an Agent-Action-Outcome narrative graph from unstructured text; Phase 2 scores it through a three-pillar verification engine (factual, causal, and evidence divergence) against four medical knowledge bases; Phase 3 computes the NTD and NRS metrics and assigns a quadrant.

### Data Sources and Corpus Characteristics

We collected 5,386 health discussion threads (185,181 comments) from four Reddit communities - r/diabetes, r/hypertension, r/loseit, and r/AskDocs, using the official Reddit API. Data spanned February 2019 to November 2024. Topic selection was informed by prior survey findings where diabetes, hypertension, and weight management were reported as the most searched health topics^4,5^. Exploratory analysis of our corpus revealed that narratives frequently contained explicit causal claims (75%), lacked source attribution (85%), and used medical terminology inconsistently (30%), motivating the three-pillar architecture.^6–8^

Previously labeled 437 reddit posts by a Dentist with Accident and Emergency (A&E) clinical experience during the COVID-19 pandemic for misinformation classification (Cohen’s κ = 0.82) provided the expert-labeled baseline NLP classifier for comparison.^7,8^ For narrative modelling, two domain experts (biomedical researcher, PhD, 25 years pharmaceutical research; medical doctor, 18 years clinical practice) independently annotated 2,000 stratified narrative segments across five dimensions: Labovian narrative stages,^11^ AAO spans, emotion intensity,^28^ epistemic stance,^41^ and BIO tags. The protocol included a calibration phase (100 segments with iterative guideline refinement and adjudication); disagreements (12.3% of annotations) were resolved through consensus adjudication, with 47 segments excluded rather than forced to resolution. Krippendorff’s α ranged from 0.78 to 0.82 across all annotation dimensions, exceeding the 0.67 threshold for acceptable reliability.^20^ The 2,000 annotated segments were drawn from 704 unique parent posts (mean 2.84 segments per narrative), which constitute the comment-level narratives used for VERITAS validation; 699 contained at least one complete agent-action-outcome triplet and were classified. The full annotation protocol is provided in Supplementary Material S3.

**Table 1.** Corpus Characteristics and Dataset Overview.

| Characteristic | Value | Details |
| --- | --- | --- |
| **Total threads** | 5,386 | From 4 subreddits (2019-2024) |
| **Total comments** | 185,181 | Average thread depth: 34.4 comments |
| **Total words** | 2,295,242 | Naturalistic health discourse |
| **Annotated segments** | 2,000 | Stratified sampling by topic/engagement |
| **Comment-level narratives** | 704 | Sufficient structure for verification |
| **Expert-labeled baseline** | 437 | Binary misinformation labels |
| **Topic: Diabetes** | 29.0% | Type 1, Type 2, medication management |
| **Topic: Obesity/Weight** | 24.9% | Diet, exercise, weight loss strategies |
| **Topic: Hypertension** | 21.9% | Blood pressure, medication adherence |
| **Topic: COVID-19** | 16.6% | Symptoms, treatments, prevention |
| **Topic: General health** | 7.7% | Multi-condition discussions |
| **Inter-rater reliability** | $\alpha=0.78-0.81$ | Krippendorff’s alpha across dimensions |

#### Phase 1: Narrative Graph Construction

Narrative graph construction, described fully in our prior work,^10^ transforms unstructured health stories into structured AAO graphs using a dual-transformer architecture. DistilBERT^16^ classifies Labovian narrative stages, emotion, stance, and tone (Labovian-stage macro-F1 = 0.501, micro-F1 = 0.609), while BioBERT^12^ with BIO sequence tagging^31^ performs biomedical entity extraction and AAO triplet identification (held-out span micro-F1 = 0.850, macro-F1 = 0.835; cross-validated span-F1 = 0.851 ± 0.007). Medical concepts are normalized to SNOMED CT using QuickUMLS and BioBERT embeddings^17,18^ (100% concept coverage via three-stage cascade: exact match, fuzzy match, and BioBERT semantic similarity). The resulting directed graphs encode agents, actions, and outcomes with temporal ordering, providing the structured input for downstream verification. Figure 2 presents an example of AAO narrative graph.

**Figure 2.**
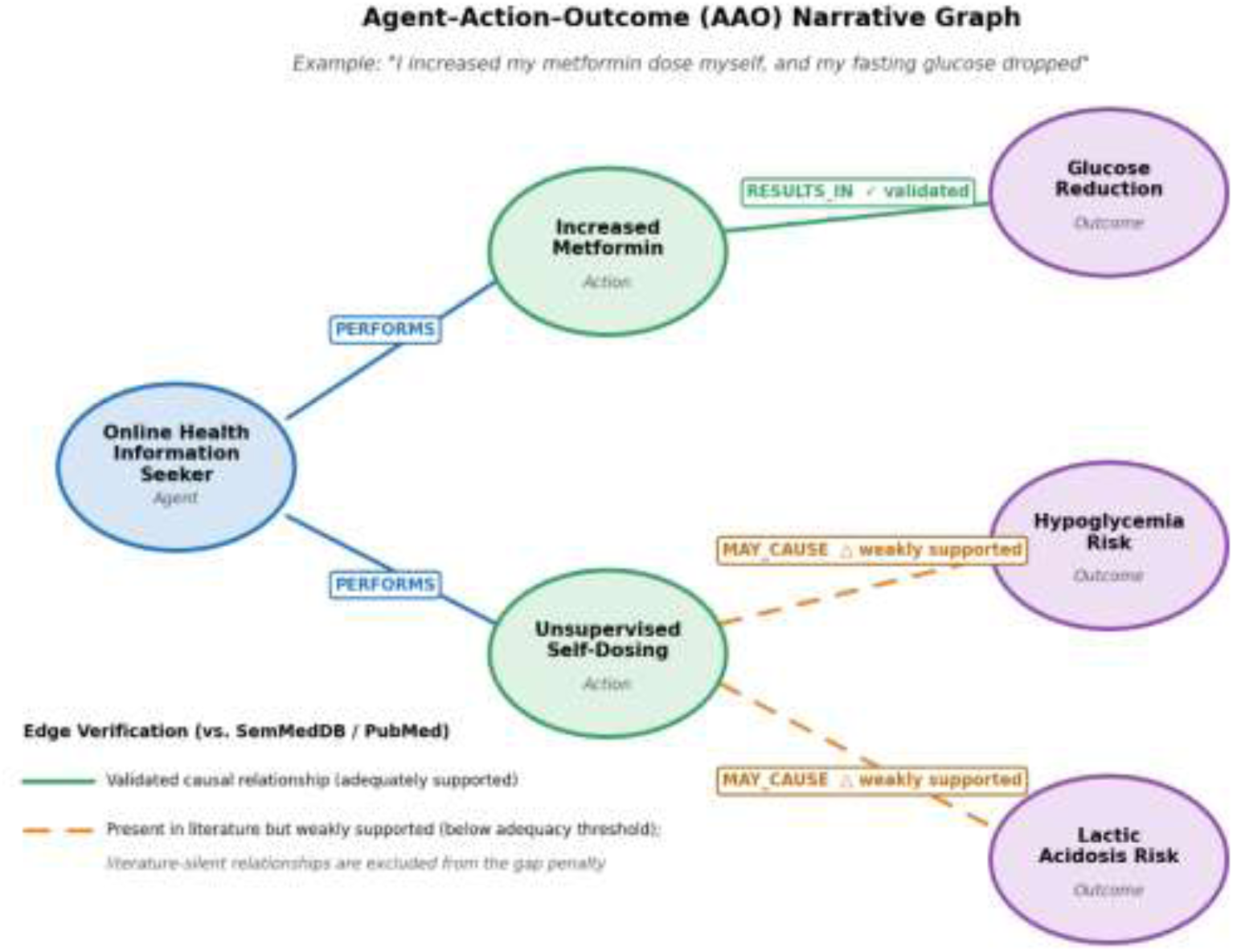
Example Agent-Action-Outcome Narrative Graph with validated and weakly supported causal relationships.

#### Phase 2: Three-Pillar Verification Engine

The AAO graphs are validated against four authoritative biomedical databases: UMLS Metathesaurus^13^ (3.45M concepts), SNOMED CT^15^ (27M concept-pair priors), SemMedDB^14^ (130M predications), and PubMed^35^ (35M publications). Epistemic divergence is computed through three complementary pillars:

**Factual Divergence (***D_F_* ) applies distributional semantics^22^ and KL divergence^26^ to measure the terminology alignment between narrative concept distributions and UMLS/SNOMED CT reference distributions.

**Causal Divergence (*D_C_*)** evaluates cause-effect plausibility against SemMedDB predications, operationalized using Pearl’s causal hierarchy^24^ to distinguish observational, interventional, and counterfactual claims. *D_C_* combines causal KL divergence, a contradiction penalty for claims opposing established evidence, and an evidence gap penalty for unsupported claims.

**Evidence Divergence (*D_E_*)** quantifies divergence from evidence-based medical research across three dimensions: content misalignment (KL divergence against PubMed topic distributions), overconfidence detection (confidence calibration theory^27^ with Wasserstein distance^34^) and evidence gap penalties. Overconfident assertions represent the primary vector for harmful misinformation^52–54^, motivating the highest component weight (0.40).

#### Phase 3: Two-Dimensional Scoring

The scoring phase measures divergence and operationalizes the Risk Irrelevance Principle through two independently computed continuous metrics.

**Narrative Truth Distance (NTD)** is a weighted combination of three divergence components, NTD = 0.25·*D_F_* + 0.35·*D_C_* + 0.40·*D_E_*, where *D_F_* (factual divergence) measures terminological departure from UMLS and SNOMED CT concepts, *D_C_* (causal divergence) measures faulty causal inference against SemMedDB predications, operationalizing Pearl’s causal hierarchy,^24^ and *D_E_* (evidence divergence) measures confidence-evidence miscalibration against PubMed.

**Narrative Risk Score (NRS)** estimates harm potential if a reader acts on the described behavior, extending Kaplan–Garrick risk formalism^25^ to online health contexts: NRS = σ(6.0·(r − 0.75)), where r = 1.0·H + 0.6·C − 0.4·M combines harm potential H (SNOMED-mapped severity, immediacy, and vulnerability, aggregated so that confluent risks outweigh isolated ones^49^), contextual amplification C, and protective mitigation M from disclaimers, consultation advice, and hedging.^43^ Component weights (α = 1.0, β = 0.6, γ = 0.4) prioritize direct clinical risk,^50,51^ and all weights and sigmoid parameters were calibrated on the annotated corpus. Complete formulations, parameter derivations, worked examples, and the weight-sensitivity analysis appear in Supplementary Material S4 and S6.

### Statistical Analysis

Independence validation employed seven methods: Pearson, Spearman, and Kendall correlations, mutual information, chi-square, logistic regression, and permutation testing (10,000 iterations). Quadrant thresholds (NTD=0.30, NRS=0.50) were optimized via ROC analysis, expert consensus, and 5-fold cross-validation. Performance comparison used McNemar’s test^44^. Shapley decomposition^18,21^ assessed component contributions.

### Quadrant Classification

AAO narrative graphs, verified through the three-pillar engine, are assigned NTD and NRS scores that jointly determine placement across four risk categories: Q1 (Aligned & Safe), Q2 (Divergent but Safe), Q3 (Aligned but Risky), and Q4 (Divergent & Risky). Under the Risk Irrelevance Principle, non-zero prevalence of Q2 and Q3 demonstrates misalignment between epistemic divergence and harm potential, revealing the structural limitations of binary classification.

## RESULTS

VERITAS is evaluated as a risk-aware classification framework, not a binary misinformation detector competing on accuracy. The primary evaluation objectives are: (1) whether NTD and NRS capture empirically distinct dimensions of online health discourse (validated through seven convergent statistical tests); (2) whether the two-dimensional quadrant decomposition reveals clinically meaningful structure that binary systems structurally cannot access; and (3) whether VERITAS improves detection coverage for content posing health risk. Expert misinformation labels serve as a comparison anchor, not as ground truth for the quadrant classification itself. These labels establish what text baselines detect versus what the two-dimensional analysis reveals. The reported precision of 29.0% reflects this deliberate broadening of scope to capture risk rather than divergence, and is a research-validation metric, not deployment-ready performance.

As a zero-shot baseline, a current-generation large language model (claude-sonnet-4-5, temperature 0) judged each of the 437 labeled posts under two single-word instructions matched to VERITAS’s axes, one asking whether acting on the post would risk harm and one asking whether it contradicts medical consensus, with three repetitions per post and majority vote.

### Dataset and Processing

VERITAS extracted 4,975 AAO triplets from 704 AAO narratives (mean 7.1 per narrative) with 100% UMLS concept coverage and mean CPU-processing time of approximately 7.2 seconds per narrative. Score distributions showed mean NTD = 0.393 (SD = 0.196) and mean NRS = 0.536 (SD = 0.131, range 0.154-0.913). Component analysis revealed moderate factual divergence (*D_F_* = 0.358), the highest divergence in causal reasoning (*D_C_* = 0.421), and moderate evidence divergence (*D_E_* = 0.391). The leading role of causal divergence is consistent with the Shapley finding that *D_C_* contributes the largest share of discriminative power.

#### Validation of Risk Irrelevance Principle

NTD and NRS satisfied the pre-specified independence criterion (|r| < 0.30) across all seven independence tests. Pearson r = 0.222 (95% CI: 0.150 to 0.291, p = 2.97e-9), Spearman ρ = 0.229 (p = 8.5e-10), and Kendall τ = 0.155 (p = 1.2e-9) confirmed a weak positive association below the practical-independence threshold. Mutual information I(D;R) = 0.096 bits fell below the 0.10 threshold, confirming weak information sharing between dimensions. Chi-square (χ² = 26.28, p = 2.9e-7, Cramér’s V = 0.194) indicated weak effect size. Logistic regression showed limited discriminative power (AUC = 0.617), and permutation testing (10,000 iterations) confirmed the observed correlation was significantly different from the null (permuted mean r = -0.0003, SD = 0.038, p < 0.0001).

**Figure 3.**
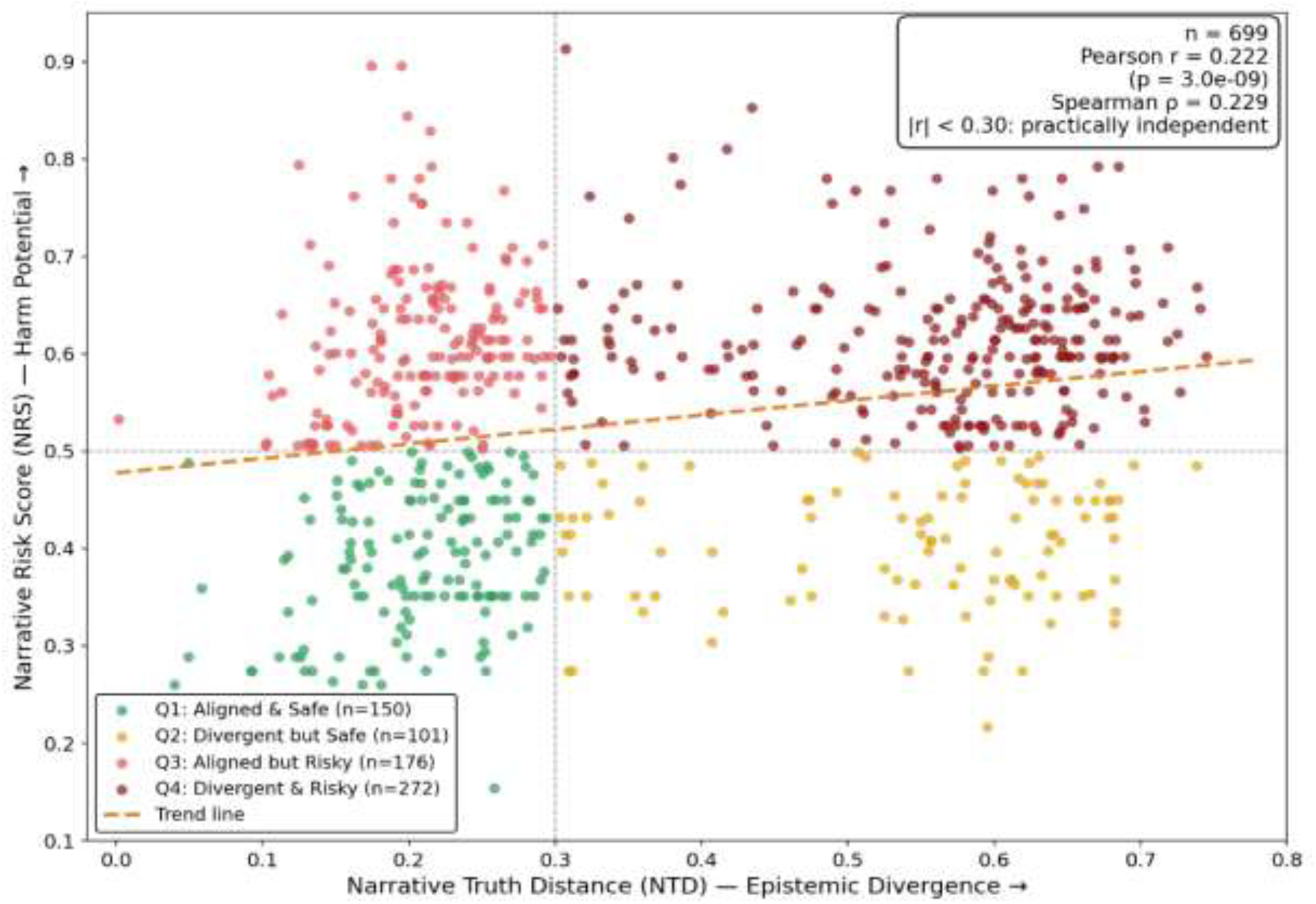
NTD-NRS Scatter Plot with Quadrant Classification. Scatter plot of the 699 classified narratives showing a weak positive association between NTD and NRS (r = +0.222, p = 3.0e-9). Dashed lines indicate the quadrant thresholds (τNTD = 0.30, τNRS = 0.50).

The association is far too weak to serve as a classification signal: divergence and risk capture substantially distinct constructs, and using divergence as a proxy for risk therefore produces systematic misclassification.^55–57^

### Quadrant Distribution

Quadrant classification revealed (N = 699 classified, 5 excluded for missing medical content): Q1 (Aligned & Safe): 150 (21.5%); Q2 (Divergent but Safe): 101 (14.4%); Q3 (Aligned but Risky): 176 (25.2%); Q4 (Divergent & Risky): 272 (38.9%). Q3 and Q4 together constitute the potentially harmful share of the corpus at 64.1%. Q3 represents an online health safety gap: content using correct medical terminology to describe unsafe self-care behaviors that binary classifiers approve as legitimate. Q2 and Q3 together constitute a structural blind spot; 39.6% of narratives systematically mishandled by binary approaches: Q3 content approved despite carrying risk, and Q2 content flagged despite being safe (Table 2).

**Table 2.** Quadrant Distribution and TF-IDF Binary Classifier Failure Analysis.

| Quadrant | Count | % | Description | Binary System View | Actual Risk | Handling |
| --- | --- | --- | --- | --- | --- | --- |
| **Q1: Aligned & Safe** | 150 | 21.5% | Low divergence, low risk. Evidence-based advice consistent with medical consensus posing minimal harm. | “Legitimate” | Safe | Correct |
| **Q2: Divergent but Safe** | 101 | 14.4% | High divergence, low risk. Traditional remedies, cultural practices, alternative approaches that diverge from consensus but pose minimal non-clinical harm. | “Misinformation” | Safe | False Positive |
| **Q3: Aligned but Risky** | 176 | 25.2% | Low divergence, high risk. Correct medical terminology used to describe potentially harmful behaviors (insulin dose adjustment, medication discontinuation, symptom dismissal). | “Legitimate” | Potentially harmful | False Negative - Critical Blind Spot |
| **Q4: Divergent & Risky** | 272 | 38.9% | High divergence, high risk. Traditional misinformation with incorrect facts AND elevated health risk. | “Misinformation” | Potentially harmful | Correct |
| **Total mishandled** | **277** | **39.6%** | **Q2+Q3: Systematic misclassification from conflating divergence with risk** | - | - | **Binary Failure Rate** |

#### Example of Aligned but Risky (Q3) Narratives

A representative Q3 narrative describes a reader doubling their own metformin dose using clinically correct terminology and plausible glucose values (Supplementary Material S5, Case Study 2): every fact is accurate, yet the unsupervised escalation carries risk that lexical verification cannot detect.

**Figure 4.**
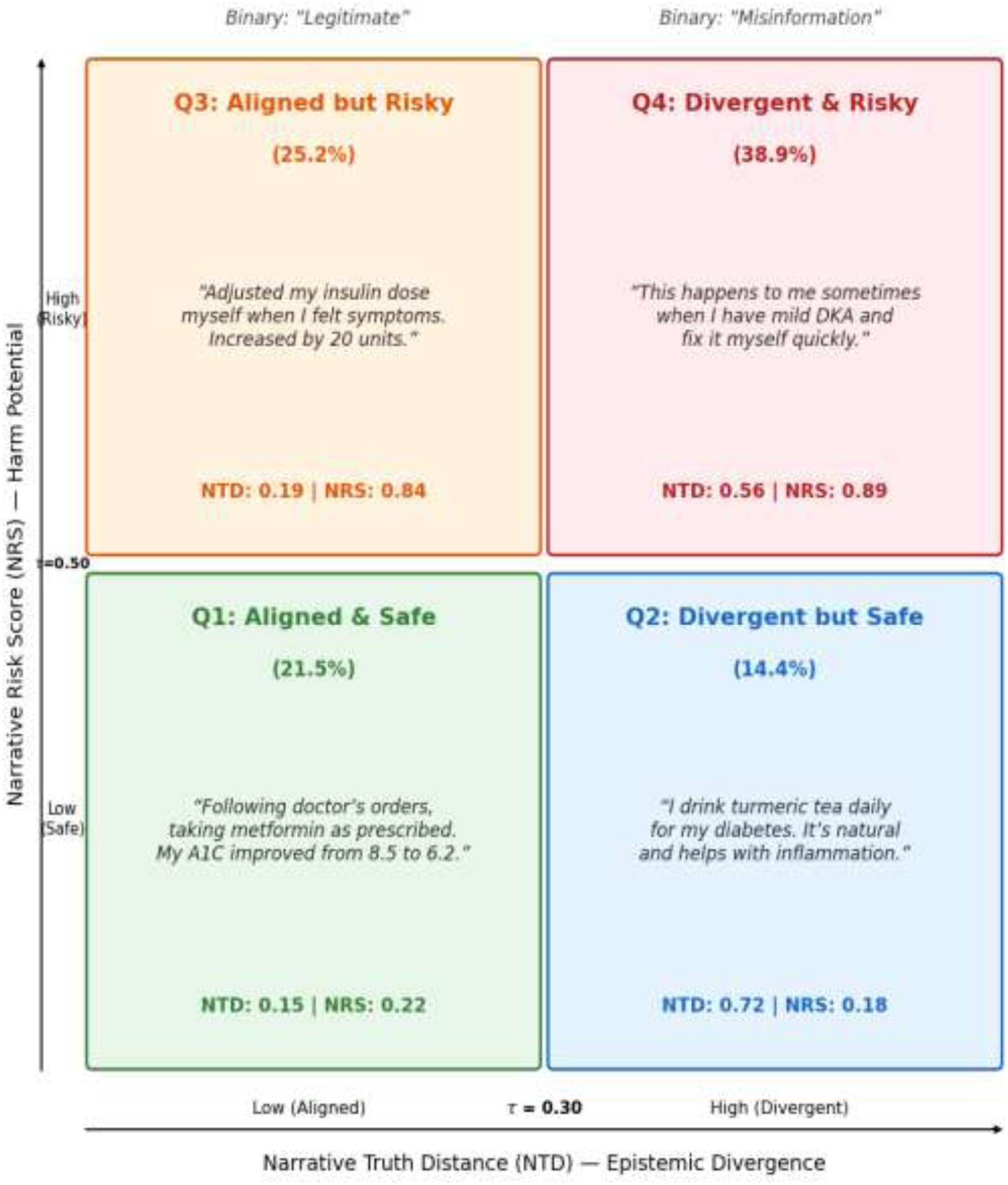
Quadrant Distribution with Representative Examples. Four VERITAS quadrants with representative narrative examples and percentage distribution across the 699 classified narratives. The Q4 panel presents an observed corpus narrative with its computed scores; the remaining panel scores are illustrative exemplars within the observed score ranges.

### Performance Comparison: VERITAS vs Base NLP Classification

In expert-labeled validation (N = 435; of the 437 expert-labeled posts, 2 lacked valid pipeline scores), VERITAS correctly identified 79 of 127 (62.2%) misinformation instances as potentially risky (Q3 or Q4) versus 73 of 127 (57.5%) for the best text baseline (BioBERT frozen embeddings with balanced class weights, stratified 5-fold cross-validation), a 4.7 percentage point improvement. Critically, 35 of 127 (27.6%) expert-labeled misinformation instances fell into Q3, the category binary classifiers systematically approve as legitimate. Combined with Q2 (n = 22, 17.3%), binary approaches mishandle 57 of 127 (44.9%) expert-labeled misinformation cases, either approving potentially harmful content (Q3) or over-flagging safe content (Q2). VERITAS classified 48 of 127 (37.8%) expert-labeled “misinformation” in safe quadrants (Q1/Q2), correctly identifying these as factually divergent but potentially unharmful.

The complete comparison across all three systems on the same expert-labeled data:

**Table 3.**
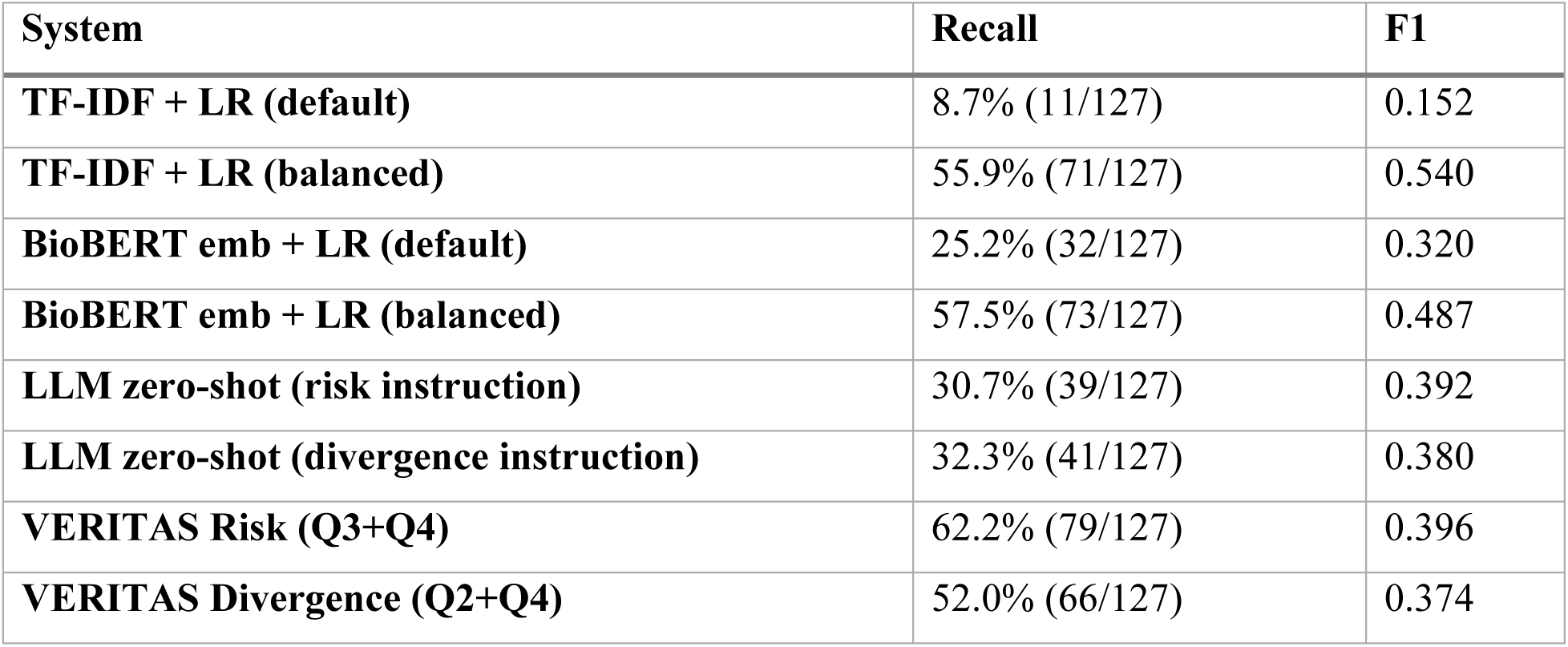
Performance Comparison: VERITAS vs Text Baselines (text baselines: stratified 5-fold cross-validation on the 437 labeled posts; VERITAS quadrant mappings: the 435 posts with complete pipeline scores; misinformation set identical, n = 127).

**Table 4.**
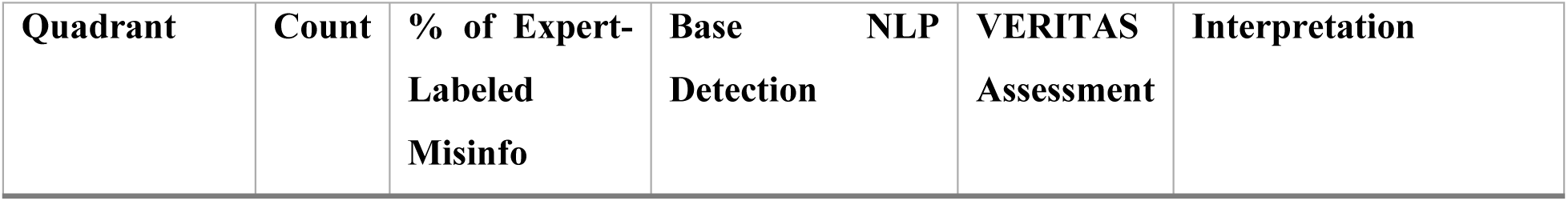
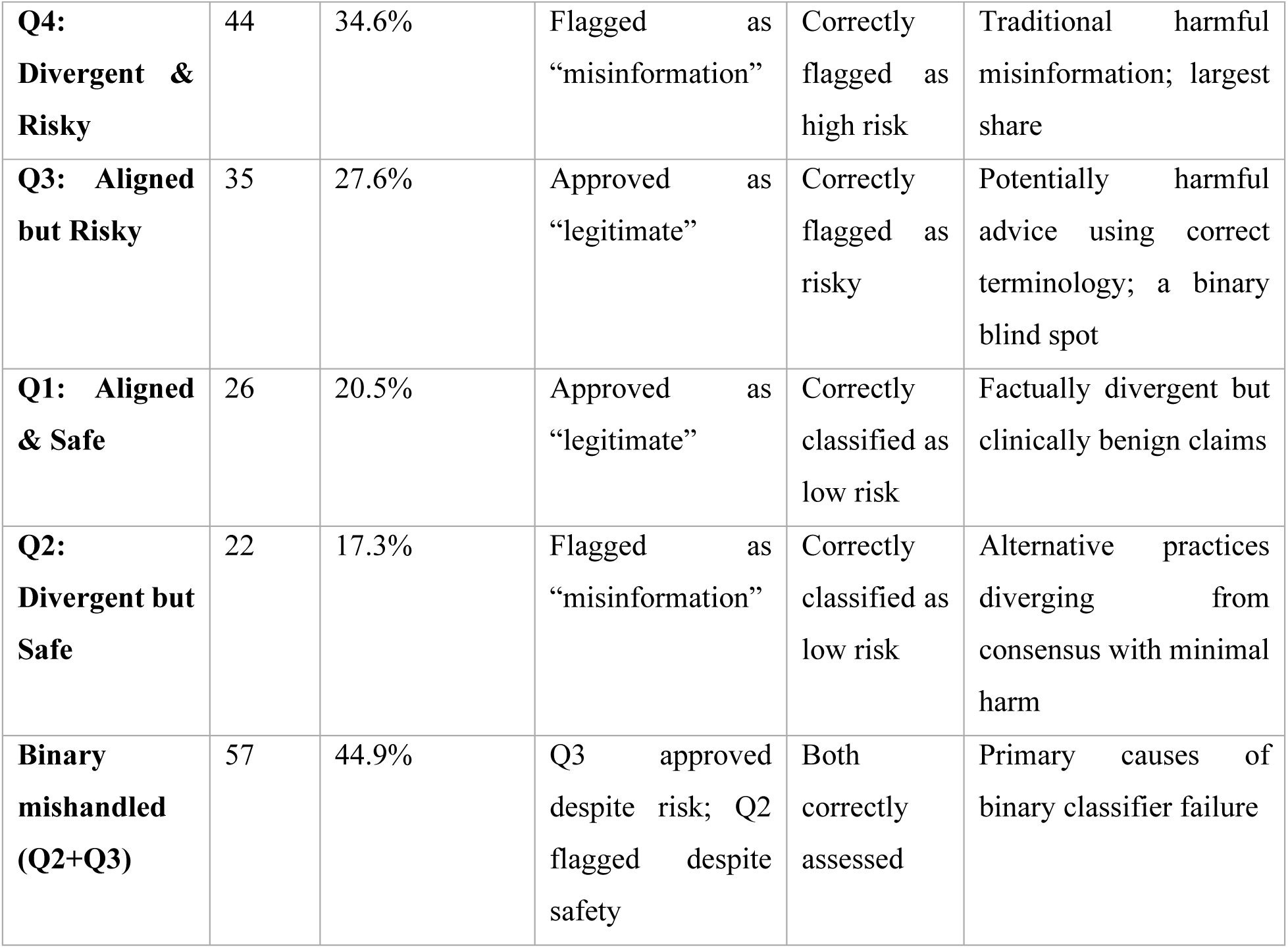
Distribution of Expert-Labeled Misinformation Across VERITAS Quadrants (n=127 expert-labeled misinformation cases). Interpretation: The largest share of expert-labeled misinformation falls into Q4 (34.6%), the divergent-and-risky category that binary classifiers can reach because these narratives are terminologically divergent. Q3 accounts for 27.6%, representing misinformation that passes binary verification because it uses correct medical terminology. Together, Q3 and Q4 (62.2%) constitute the potentially harmful misinformation that VERITAS risk mapping detects. The 37.8% reclassified into safe quadrants (Q1 and Q2) reflects content that diverges from consensus without posing actionable health risk.

| Quadrant | Count | % of Expert-Labeled Misinfo | Base NLP Detection | VERITAS Assessment | Interpretation |
| --- | --- | --- | --- | --- | --- |
| **Q4: Divergent & Risky** | 44 | 34.6% | Flagged as “misinformation” | Correctly flagged as high risk | Traditional harmful misinformation; largest share |
| **Q3: Aligned but Risky** | 35 | 27.6% | Approved as “legitimate” | Correctly flagged as risky | Potentially harmful advice using correct terminology; a binary blind spot |
| **Q1: Aligned & Safe** | 26 | 20.5% | Approved as “legitimate” | Correctly classified as low risk | Factually divergent but clinically benign claims |
| **Q2: Divergent but Safe** | 22 | 17.3% | Flagged as “misinformation” | Correctly classified as low risk | Alternative practices diverging from consensus with minimal harm |
| **Binary mishandled (Q2+Q3)** | 57 | 44.9% | Q3 approved despite risk; Q2 flagged despite safety | Both correctly assessed | Primary causes of binary classifier failure |

The two quadrant mappings share a common core by construction: both flag the 44 Q4 narratives (34.6%) that are simultaneously divergent and risky, matching the traditional conception of misinformation. Their difference is a structural exchange of one off-diagonal quadrant for the other. The divergence mapping adds the 22 Q2 narratives (17.3%) that diverge from consensus yet carry little harm potential (52.0%, 66/127); the risk mapping instead adds the 35 Q3 narratives (27.6%) that are epistemically aligned yet carry elevated harm potential (62.2%, 79/127). The recall difference thus reflects an exchange of error types, over-flagged safe content for recovered aligned-but-risky content, rather than incremental tuning.

A zero-shot large language model (claude-sonnet-4-5, temperature 0) recovered 30.7% (39/127) of expert-labeled misinformation under the risk instruction and 32.3% (41/127) under the divergence instruction, against 62.2% for VERITAS’s risk mapping on the same misinformation set. The model operated at a conservative point (precision 54.2% versus VERITAS’s 29.0%) with comparable F1 (0.392 versus 0.396); under the safety framing in which missed harmful content is the costly error, the recall gap is the operative comparison.

**Figure 5.**
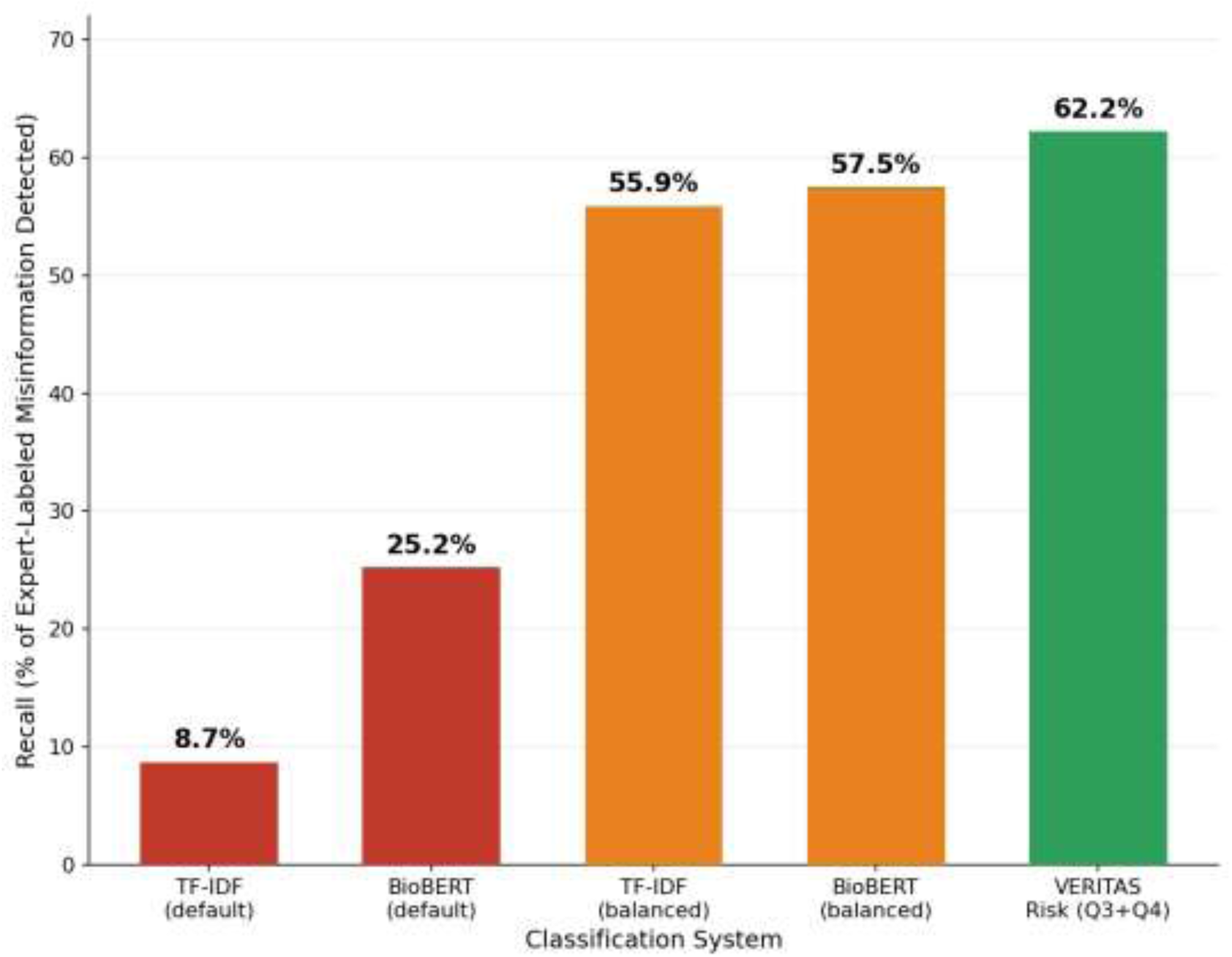
VERITAS vs Text Baseline Classification Performance. Recall comparison: VERITAS risk mapping (62.2%) versus best text baseline (BioBERT balanced, 57.5%); default and class-weight-balanced configurations are shown for both text baselines. The risk axis recovers misinformation that text classifiers miss by measuring behavioral harm potential rather than lexical divergence.

#### Component Contribution Analysis

Ablation analysis confirms that all three NTD components contribute meaningfully. Shapley value decomposition attributed discriminative power as: *D_C_* (Causal) 39.7%, *D_E_* (Evidence) 35.3%, *D_F_* (Factual) 25.0%. The causal component’s leading contribution confirms the theoretical prediction that health misinformation reflects primarily faulty inference rather than terminology errors. VERITAS’s causal direction detection, distinguishing ‘exercise causes glucose reduction’ (supported) from ‘glucose reduction causes exercise initiation’ (implausible inversion), corrected 346 segment-level causal inversions (137 narratives exhibit reactive outcome-to-action causation). Weight sensitivity analysis across 11 configurations confirmed that all maintained |r| < 0.30, demonstrating a weak association below the practical-independence threshold, robust to parameter variation.

**Figure 6.**
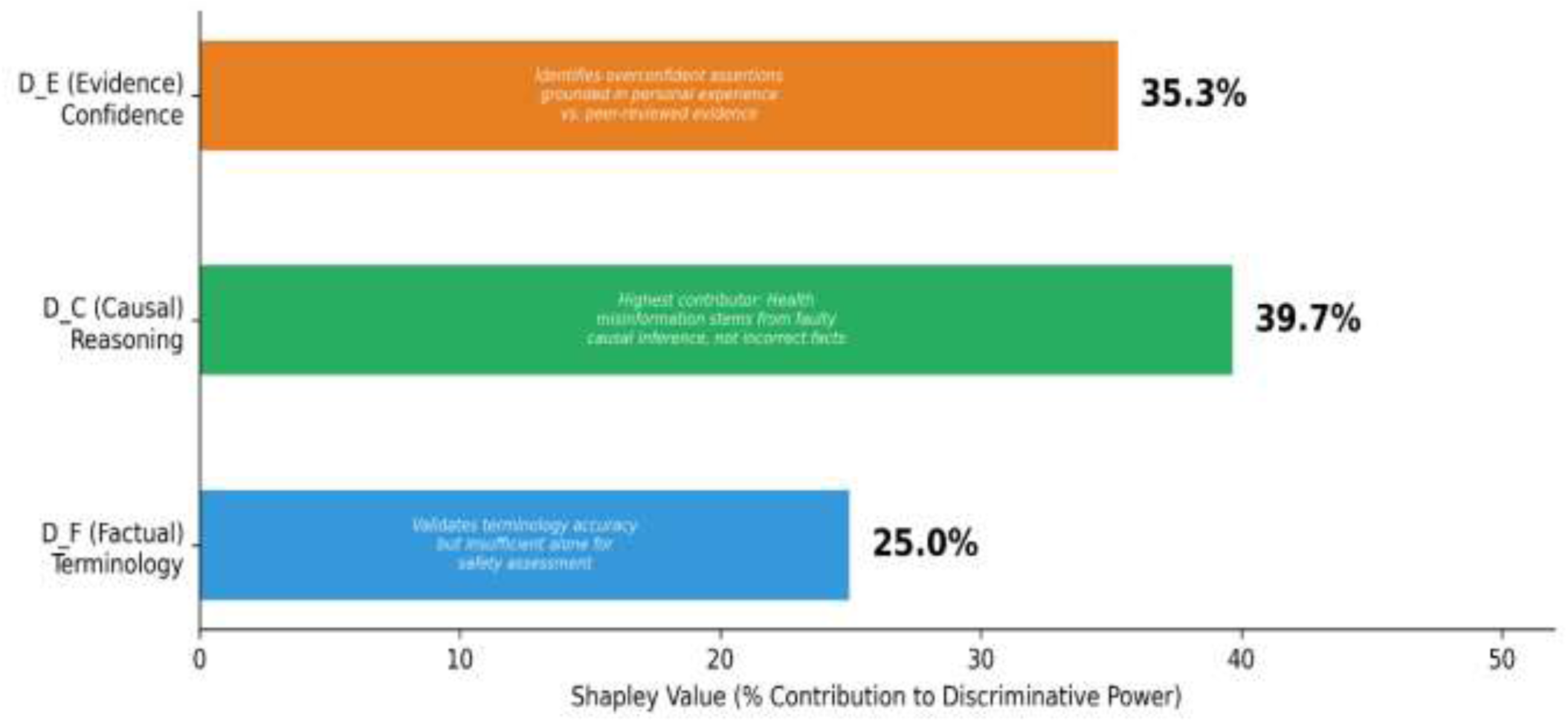
Shapley Value Component Analysis. Shapley value decomposition revealing discriminative power attribution across VERITAS’s three epistemic divergence components. Causal Divergence (D_C_) contributes 39.7% to discriminative power, validating that health misinformation primarily reflects faulty causal inference rather than terminology errors or overconfidence alone.

## DISCUSSION

For individuals guiding self-care using peer-generated health content, the central finding is that terminological correctness provides no reliable signal of behavioral safety: whether a post uses accurate medical language says little about whether acting on it is safe, a gap readers without clinical training cannot see past. Seven independence tests confirm this: epistemic divergence and harm potential are only weakly associated (r = 0.222, 4.9% shared variance), robust across all 11 weight configurations. The corpus mean NRS (0.536) reflects the chronic disease self-management focus of the sampled Reddit communities, where medication adjustment and treatment modification are disproportionately represented; this compositional bias should be considered when generalizing Q3 prevalence to broader online health discourse.

The potentially harmful share (Q3 + Q4 = 64.1%) demonstrates that the risk-divergence misalignment is substantial, not a marginal edge case. Q3 at 25.2% and Q2 at 17.3% represent structural blind spots, content that is either terminologically aligned yet behaviorally risky or divergent from consensus but poses minimal harm, invisible to any single-axis system. Moderate factual divergence (*D_F_* = 0.358) confirms that risky narratives share the medical vocabulary of safe content, the Lycett-Partridge ontology-epistemology distinction operationalized empirically^36^. The 39.7% Shapley contribution of causal divergence (*D_C_*)) reveals that potentially harmful advice stems from inappropriate causal inferences using accurate terminology, a pattern lexical verification cannot detect^6,7^. The 346 segment-level causal inversions corrected by VERITAS provide concrete evidence that the dominant failure mode is reasoning structure rather than factual content. These findings align with recent systematic reviews demonstrating that content-based detection methods remain limited by reliance on surface linguistic features rather than reasoning structure^45,46^.

The 37.8% reclassification finding (48/127), where VERITAS placed expert-labeled “misinformation” in safe quadrants (Q1/Q2), indicates that the framework’s risk dimension diverges from binary expert judgment on a substantial minority of cases. We interpret this cautiously and as hypothesis-generating rather than corrective: whether these narratives are genuinely low-risk requires external clinical validation, which our preliminary expert review has begun but not yet established at scale. If borne out, the pattern would suggest that binary expert annotation can conflate epistemic divergence with harm, the same limitation exhibited by automated classifiers, with implications for how ground-truth labels are constructed in future health misinformation datasets.

Zero-shot single-word prompting represents a lower bound on large language model capability rather than a ceiling; few-shot or chain-of-thought prompting would likely raise recall. The comparison establishes what off-the-shelf instruction-following achieves under axis-matched prompts, and VERITAS’s advantage additionally includes component-level, auditable scores (*D_F_*, *D_C_*, *D_E_*, NTD, NRS) rather than an unexplained verdict.

VERITAS achieves a 4.7-point recall gain over the best text baseline (62.2% vs 57.5%) at a precision cost (29.0% vs 42.2%). This trade-off reflects the asymmetric error cost structure governing online health safety: a false negative (approving potentially harmful content) carries greater consequence than a false positive (flagging safe content for review). Binary classification concentrates detections on lexically divergent posts (Q2/Q4), missing the 27.6% of misinformation using correct medical terminology (Q3). The independent NRS dimension identifies the Q3 category by measuring behavioral risk rather than lexical divergence, recovering a category of potentially harmful content that single-axis systems structurally cannot reach.

### Implications for Online Health Safety Verification

These findings reframe the computational challenge of online health safety. The dominant paradigm organizes verification around factual accuracy, treating correctness as a proxy for safety. The Risk Irrelevance Principle shows this framing is inadequate: truth-conditional classification captures only one dimension of a two-dimensional problem, systematically missing the Q3 category (25.2% of the corpus) where factual fluency masks behavioral risk. The phenomenon parallels what Marwick and Boyd^48^ term “context collapse”: just as platform architectures flatten distinct audiences into a single context, binary classifiers flatten distinct epistemic dimensions into a single verdict.

This has implications at three governance levels. At the platform level, Q3 narratives can receive contextual safety warnings rather than removal, preserving informational access while flagging health risk. At the clinical level, patient portals could integrate narrative risk signals to detect unsupervised medication changes or delayed care-seeking. At the public health level, population-scale narrative risk surveillance could identify emerging unsafe self-care patterns before they manifest as adverse outcomes, complementing existing pharmacovigilance systems. At a mean 7.2 seconds per narrative, VERITAS operates within feasible range for near-real-time integration.

The NRS bands translate directly into clinical action (Supplementary Material S4, Table S4.2): scores at or above 0.60 flag content for expert review, and the graduated response ranges from contextualization (Q2) through warning (Q3) to restriction (Q4).

### Health Equity Implications

The Risk Irrelevance Principle has significant equity implications. Accuracy-driven moderation disproportionately flags alternative, traditional, or culturally grounded health practices while missing biomedically fluent but unsafe advice, reinforcing cultural bias in content governance.^1,3–5^ These dynamics particularly affect communities relying on non-Western health knowledge. Decoupling divergence from risk enables proportionate moderation; low-risk divergent practices (Q2) are contextualized rather than censored, while high-risk aligned narratives (Q3) receive appropriate safety warnings regardless of terminological correctness.

#### Limitations

Several limitations apply. First, UMLS concept normalization, while achieving 100% coverage, may not capture emerging terminology, colloquial drug names, or culturally specific health concepts. Second, sigmoid parameters (scale=6.0, center=0.75) and quadrant thresholds were calibrated on a single corpus; external validation across diverse populations and platforms is needed. Third, annotation relied on two expert reviewers, and the comparison against zero-shot binary baselines, while demonstrating structural limitations, does not represent the strongest possible binary classifier.⁴⁵ Fourth, the corpus is drawn from a single platform (Reddit) and limited to English-language content from 2019-2024, and may not generalize to other online health settings. Implementation challenges including alert fatigue, privacy concerns, and liability considerations remain substantial; however, given the morbidity from medication non-adherence³² and inappropriate self-modification, even modest improvements in detecting unsafe behaviors could yield meaningful health benefit.

#### Future Directions

Future work should evaluate VERITAS across additional platforms, incorporate multimodal narratives, extend to non-English contexts, and support real-time deployment with dynamic knowledge base updates. Formalizing Risk Irrelevance enables development of provably risk-aware systems with explicit guarantees about failure modes.

#### Conclusion

Epistemic divergence and harm potential are weakly associated dimensions requiring separate assessment. Across 704 health narratives (699 classified), seven independence tests confirm this structural property (Pearson r = 0.222, I(D;R) = 0.096 bits), demonstrating that conflating factual divergence with harm potential is both theoretically unsound and practically costly for online health safety. By computing Narrative Truth Distance and Narrative Risk Score independently, VERITAS exposes the two categories of unsafe health content: Q4 (divergent and risky, 38.9%), which binary classifiers can reach, and Q3 (aligned but risky, 25.2%), advice that is terminologically correct yet potentially harmful, which no single-axis system can detect. The prominence of causal reasoning errors (*D_C_* Shapley contribution 39.7%) confirms that online health misinformation is primarily a problem of faulty inference rather than incorrect facts. The recall advantage over single-axis detection is therefore best understood not as an incremental improvement but as an exchange of error types: over-flagged divergent-but-safe content is released while aligned-but-risky content, invisible to divergence-based screening, is recovered. As online health information increasingly shapes self-care decisions between clinical encounters, the ability to distinguish what is cultural or alternative from what is potentially harmful is essential for platform governance and for ensuring that individuals have the means to verify that the information they encounter is safe to act upon.

## Acknowledgments

The authors thank the clinical domain experts who contributed to annotation development and validation of narrative graphs. We acknowledge the computational resources provided by the University of Maryland, Baltimore County.

## Authors’ Contributions

Following CRediT (Contributor Roles Taxonomy):

**OC (Ommo Clark):** Conceptualization, Methodology, Software, Validation, Formal Analysis, Investigation, Data Curation, Writing - Original Draft, Writing - Review & Editing, Visualization. **AJ (Anupam Joshi):** Conceptualization, Methodology, Resources, Writing - Review & Editing. **KPJ (Karuna P. Joshi):** Conceptualization, Resources, Writing - Review & Editing, Supervision, Project Administration.

## Ethics and Data Governance

This study uses publicly available Reddit data collected via official API in compliance with Reddit Terms of Service and established guidelines for social media research ethics.63,64 No personally identifiable information was collected. The study was determined exempt from Institutional Review Board review under 45 CFR 46.104(d)(4), consistent with research involving publicly available data which covers secondary research use of publicly available information. All narrative examples in this manuscript have been paraphrased to prevent re-identification.

## Supplementary Material

- S1. Data_collection_methods.pdf - Reddit data collection protocol for corpus replication
- S2. database_access_guide.pdf - Medical knowledge base access instructions
- S3. annotation_guidelines.pdf - Complete expert annotation protocol Additional Materials:
- S4. mathematical_appendix.pdf - Complete NTD and NRS formulations with worked examples S5. extended_examples.pdf - Detailed narrative case studies by quadrant with AAO graphs
- S6. sensitivity_analysis.pdf - Weight optimization and robustness validation

## Data Availability

### Code and Algorithms

The complete VERITAS source code, including the AAO extraction pipeline (aao_pipeline_production_v3_4.py), NTD/NRS computation modules (ntd_nrs_quadrant_classifier_v3_4.py, nrs_batch_calculator_v5.py), individual divergence calculators (df_calculator_v3_4.py, dc_calculator_v3_4.py, de_calculator_v3_4.py), and all experiment scripts (ablation, weight sensitivity, baseline comparison), will be released within 30 days of manuscript acceptance with permanent archiving at the KnACC Lab, UMBC (DOI to be assigned).

### Data Sources

This study used publicly available Reddit data collected via official API. Due to Reddit Terms of Service restrictions and user privacy considerations, the corpus cannot be redistributed.

### Resources

UMLS, SNOMED CT, SemMedDB, and PubMed are publicly available with separate licensing. Access instructions are provided in Supplementary Material S2.

## Use of Generative AI

Portions of the manuscript were edited and streamlined for length and clarity using a generative language model (Claude, Anthropic). AI assistance was limited to prose revision and structural reorganization to meet journal formatting requirements; it was not used for data analysis, statistical computation, or interpretation of results. All scientific content, analyses, and interpretations were authored and verified by the human authors, who reviewed and approved all AI-assisted edits and take full responsibility for the content.

