## Supplementary File 4 for "VERITAS: A Neuro-Symbolic Approach to Quantifying Epistemic Divergence and Harm Potential in Online Health Narratives"

### Complete NTD and NRS Formulations with Worked Examples

#### 1. Theoretical Foundations

The VERITAS metrics are grounded in six theoretical frameworks: Distributional Semantics (Harris, 1954), Information Theory (Kullback & Leibler, 1951; Shannon, 1948), Causal Reasoning (Pearl, 2000), Statistical Epistemic Logic (Halpern, 2003), Optimal Transport Theory (Villani, 2009), and Risk Assessment Theory (Kaplan & Garrick, 1981).

#### 2. Narrative Truth Distance (NTD)

```
NTD = 0.25 × D_F + 0.35 × D_C + 0.40 × D_E
```

ranging [0, 1], where 0 indicates perfect consensus alignment.

##### 2.1 Factual Divergence (D_F)

```
D_F = 1 − exp(−D_KL(P_narrative || P_reference))
```

Worked example: a diabetes narrative using metformin, glucose, and A1C concepts consistently with reference usage. D_KL = 0.06, so D_F = 1 − exp(−0.06) = 0.058.

##### 2.2 Causal Divergence (D_C)

```
D_C = 1 − exp(−(D_KL_causal + 0.5 × Σκ_ij + 0.3 × N_gap))
```

Worked example: “skipping insulin when feeling fine” contradicts the evidence base. With D_KL_causal = 0.1 and one strong contradiction ($\kappa = 1.0$): D_C = 1 − exp(−0.6) = 0.451.

##### 2.3 Evidence Divergence (D_E)

```
D_E = 0.40 × E_conf + 0.35 × E_content + 0.25 × E_gap
```

Worked example: “Metformin 2000 mg is perfectly safe.” E_conf = 0.45, E_content = 0.15, E_gap = 0.30. D_E = 0.308.

#### 3. Narrative Risk Score (NRS)

```
NRS = 1 / (1 + exp(−6.0 × (r − 0.75))),   r = 1.0 × H + 0.6 × C − 0.4 × M
```

##### 3.1 Harm Potential (H)

$$H = (H\_severity \times H\_immediacy \times H\_vulnerability)^{1/3}$$

The geometric mean ensures all dimensions contribute.

*Table S4.1. Harm potential scales.*

| Component | Scale | Values |
|---|---|---|
| H_severity | SNOMED CT-mapped outcomes | 0.0 (none) to 1.0 (death) |
| H_immediacy | Temporal urgency | 1.0 (immediate) to 0.0 (months) |
| H_vulnerability | Population risk | 1.0 (critical) to 0.0 (general) |

##### 3.2 Amplification (C) and Mitigation (M)

```
C = 0.4 × C_virality + 0.4 × C_certainty + 0.2 × C_emotional
M = 0.4 × M_disclaimer + 0.4 × M_consult + 0.2 × M_hedging
```

##### 3.3 Worked Example: Complete NRS

Metformin self-dosing: H_severity = 0.6, H_immediacy = 0.5, H_vulnerability = 0.5. H = $(0.6 \times 0.5 \times 0.5)^{1/3}$ = 0.531. C = 0.44, M = 0.0. r = 1.0 × 0.531 + 0.6 × 0.44 = 0.795. NRS = 1 / (1 + exp(−6.0 × (0.795 − 0.75))) = 1 / (1 + exp(−0.27)) = 0.567 (moderate-high risk).

#### 4. Clinical Interpretation

*Table S4.2. Clinical interpretation thresholds.*

| Metric | Range | Interpretation | Action |
|---|---|---|---|
| NTD | < 0.20 | Consensus-aligned | No intervention |
| NTD | 0.20–0.40 | Caution | Contextual information |
| NTD | 0.40–0.60 | Skepticism | Active verification |
| NTD | ≥ 0.60 | Serious divergence | Professional consult |
| NRS | < 0.20 | Minimal | No intervention |
| NRS | 0.20–0.39 | Low | Monitoring |
| NRS | 0.40–0.59 | Moderate | Health education |
| NRS | 0.60–0.79 | High | Clinical intervention |
| NRS | ≥ 0.80 | Critical | Urgent intervention |

#### 5. Quadrant Classification

*Table S4.3. VERITAS quadrant framework.*

| Quadrant | Criteria | Binary System View | Risk-Aware Assessment |
|---|---|---|---|
| Q1: Aligned & Safe | NTD < 0.30, NRS < 0.50 | Legitimate | Low harm potential |
| Q2: Divergent but Safe | NTD ≥ 0.30, NRS < 0.50 | Misinformation | Low harm potential |
| Q3: Aligned but Risky | NTD < 0.30, NRS ≥ 0.50 | Legitimate | Elevated harm potential (the safety gap) |
| Q4: Divergent & Risky | NTD ≥ 0.30, NRS ≥ 0.50 | Misinformation | Elevated harm potential |
