## Supplementary File 3 for "VERITAS: A Neuro-Symbolic Approach to Quantifying Epistemic Divergence and Harm Potential in Online Health Narratives"

### Complete Expert Annotation Protocol

#### 1. Annotator Qualifications

**Annotator 1 (Biomedical Expert):** PhD in Neuroimmunology; 25 years of pharmaceutical research. Primary responsibility: medical accuracy, entity identification, UMLS concept validation.

**Annotator 2 (Clinical Expert):** MD; 18 years of clinical practice; board-certified general practitioner and OB/GYN specialist. Primary responsibility: clinical plausibility, causal assessment, clinical context.

#### 2. Training and Calibration

**Phase 1 (2 hours):** review of frameworks (Labov, Plutchik, Horne, Street); study of definitions; review of BIO tagging; practice with 10 examples. **Phase 2 (n = 100):** independent calibration with weekly meetings; target α ≥ 0.75. **Phase 3 (n = 2,000):** independent annotation with weekly check-ins; ambiguous cases flagged for adjudication.

#### 3. Labovian Narrative Stages

*Table S3.1. Labovian narrative stage definitions (Labov & Waletzky, 1967; Labov, 1972).*

| Stage | Definition | Example |
|---|---|---|
| Abstract | Summary overview | I had a scary hypoglycemic episode |
| Orientation | Contextual grounding | I'm 26, diabetic for 3 years, on metformin |
| Complicating Action | Core event sequence | My blood sugar dropped to 2.4 mmol/L |
| Evaluation | Narrator's interpretation | It was the scariest moment of my life |
| Resolution | Outcome/consequence | The ER gave me glucose and I stabilized |
| Coda | Return to present | Now I always carry glucose tablets |

#### 4. Interpretive Dimensions

##### 4.1 Emotion (Plutchik, 1980)

| Label | Definition | Example |
|---|---|---|
| Fear | Anxiety about diagnosis/treatment | Terrified about what the doctor might find |
| Hope | Optimism about recovery | I believe this new medication will help |
| Frustration | Irritation with failures | Tried everything and nothing works |
| Relief | Comfort after improvement | Test came back negative, I can breathe |
| Anger | Resentment toward providers | My doctor dismissed my concerns |

##### 4.2 Stance (Horne, 1999; DiMatteo, 2004)

| Label | Definition | Example |
|---|---|---|
| Compliant | Follows medical advice | I take medication exactly as prescribed |
| Resistant | Opposes/modifies advice | I stopped the pills, they made me worse |
| Questioning | Expresses uncertainty | Should I be on this high a dose? |
| Advocating | Promotes treatments | You should definitely try keto |
| Neutral | Reports without evaluation | Started metformin, A1C is now 6.5 |

##### 4.3 Tone (Street, 2009)

| Label | Definition | Example |
|---|---|---|
| Clinical | Objective, medical register | Fasting glucose 8.2 mmol/L; commenced metformin |
| Anxious | Worry, seeks reassurance | Really scared, what if something goes wrong? |
| Confident | Asserts with certainty | Trust me, here's what you need to do |
| Supportive | Encouragement, empathy | You've got this! We're all here for you |
| Skeptical | Doubt about treatments | Big Pharma just wants your money |

#### 5. BIO Tagging Schema

*Table S3.5. BIO tagging for agent-action-outcome (AAO) extraction.*

| Tag | Component | Description | Examples |
|---|---|---|---|
| B/I-AGENT | Agent | Who performs/experiences | I, my doctor, the nurse |
| B/I-ACTION | Action | Interventions, behaviors | started metformin, skipped dose |
| B/I-OUTCOME | Outcome | Results, consequences | blood sugar dropped, felt better |
| B/I-QUANT | Quantifier | Clinical measurements | 2.4 mmol/L, 140/90 mmHg |
| O | Outside | Not an AAO component | Connectives, prepositions |

#### 6. Edge Case Handling

**Ambiguous agency:** default to the most plausible actor. **Multiple emotions:** prioritize the primary emotion; flag "Mixed" for adjudication. **Overlapping functions:** dominant-function approach. **Implicit causal links:** mark action/outcome even without a connective, with a 50-token proximity default. **Non-narrative:** mark as non-narrative; only segments with at least one agent, one action, and one outcome proceed.

#### 7. Inter-Rater Reliability

*Table S3.6. Krippendorff's α reliability by dimension. Interpretive dimensions (Labovian stage, emotion, stance, tone) range α = 0.78–0.81; AAO span tagging ranges α = 0.75–0.82.*

| Dimension | Krippendorff's α | Agreement | Interpretation |
|---|---|---|---|
| Labovian stages | 0.81 | 88.9% | Excellent |
| Emotion | 0.79 | 87.2% | Good |
| Stance | 0.78 | 86.5% | Good |
| Tone | 0.78 | 86.3% | Good |
| AAO agents | 0.82 | 89.4% | Excellent |
| AAO actions | 0.77 | 85.9% | Good |
| AAO outcomes | 0.75 | 84.6% | Good |

Total disagreements: 246/2,000 (12.3%). Breakdown: stage boundaries (92, 37.4%), emotion ambiguity (68, 27.6%), stance interpretation (51, 20.7%), and agent identification (35, 14.2%). All were resolved by consensus.
