## Supplementary File 2 for "VERITAS: A Neuro-Symbolic Approach to Quantifying Epistemic Divergence and Harm Potential in Online Health Narratives"

### Medical Knowledge Base Access Instructions

#### 1. Overview

VERITAS validates health narratives against four authoritative medical knowledge bases through its three-pillar verification engine. This document provides access instructions, licensing requirements, and configuration details for each database.

*Table S2.1. Medical knowledge bases used in VERITAS.*

| Database | Version | Size | VERITAS Role | License |
|---|---|---|---|---|
| UMLS Metathesaurus | 2024AA | 3.45M concepts | Terminology (D_F) | Free (UMLS) |
| SNOMED CT | Jan 2024 | 27M concept pairs | Semantics (D_F, D_C) | Free (NLM) |
| SemMedDB | v43.0 | 130M+ predications | Causal (D_C) | Free (NLM) |
| PubMed/MEDLINE | 2024 baseline | 35M+ publications | Evidence (D_E) | Free (NCBI) |

#### 2. UMLS Metathesaurus

The Unified Medical Language System (UMLS) Metathesaurus integrates biomedical terminology from over 200 source vocabularies (Bodenreider, 2004). Access requires a free UMLS Terminology Services (UTS) account. Registration: https://uts.nlm.nih.gov/uts/. Key files: MRCONSO.RRF (concept names and synonyms), MRREL.RRF (relationships), MRSTY.RRF (semantic types). VERITAS uses a hybrid medical concept normalizer combining SQLite string matching (threshold = 0.7) and BioBERT semantic matching (threshold = 0.6) using precomputed embeddings (~0.6 GB), achieving 100% concept coverage on the annotated corpus via a three-stage cascade (exact match, fuzzy match, semantic match).

#### 3. SNOMED CT

SNOMED CT provides clinical terminology for semantic relationship validation. Available free through the UMLS license or from SNOMED International (https://www.snomed.org/). Key components: concept hierarchy, finding-site relationships, causative-agent relationships, and associated-morphology relationships. VERITAS uses 27 million concept-pair priors.

#### 4. SemMedDB

SemMedDB contains subject-predicate-object triples extracted from PubMed abstracts using SemRep (Kilicoglu et al., 2012). Access: https://lhncbc.nlm.nih.gov/ii/tools/SemRep_SemMedDB_SKR/SemMedDB.html. Format: MySQL dump files, which VERITAS converts to SQLite. Key tables: PREDICATION, SENTENCE, CITATIONS. VERITAS queries using UMLS CUI pairs, matching against 63 predicate types to validate causal claims.

#### 5. PubMed API

PubMed provides the evidence base for D_E computation. API access: NCBI E-utilities (https://www.ncbi.nlm.nih.gov/books/NBK25501/). Rate limits: 10 requests/second with an API key. VERITAS uses PubMed to verify claims, compute content misalignment via KL divergence, detect overconfidence, and identify evidence gaps.

#### 6. Local Database Setup

For reproducibility, VERITAS operates with local SQLite copies: cui-semtypes.db (UMLS mappings), hybrid_filtered_concepts.pkl (BioBERT embeddings, ~0.6 GB), semmeddb_local.db (SemMedDB predications), and snomed_relations.db (SNOMED CT relationships).
