## Supplementary File 1 for "VERITAS: A Neuro-Symbolic Approach to Quantifying Epistemic Divergence and Harm Potential in Online Health Narratives"

### Reddit Data Collection Protocol: Corpus Replication Guide

#### 1. Overview

This document provides the complete data collection protocol for replicating the Reddit health narrative corpus used in the VERITAS framework evaluation. The corpus comprises 5,386 health discussion threads containing 185,181 comments (2,295,242 total words) collected from four Reddit communities between February 2019 and November 2024. Topic selection was informed by prior survey findings (Clark et al., ACM SaT-CPS 2024; Clark et al., AMIA 2024) identifying diabetes, hypertension, and weight management as the most commonly searched health topics requiring frequent self-management decisions between clinical encounters.

#### 2. Target Subreddit Communities

Reddit was selected based on established criteria for social media health research. The platform's pseudonymous structure encourages candid health disclosure, its long-form conversational style supports narrative-rich content (mean thread length: 427 words), community-based moderation provides baseline quality control, and publicly accessible data via the official API enables compliant data collection.

*Table S1.1. Target subreddit communities and selection rationale.*

| Subreddit | Topic Focus | Members | Selection Rationale |
|---|---|---|---|
| r/diabetes | Type 1 & 2 diabetes | >200,000 | Most searched chronic condition |
| r/hypertension | Blood pressure management | >50,000 | Second most searched |
| r/loseit | Weight loss/management | >3,000,000 | Third most searched |
| r/AskDocs | General health questions | >1,000,000 | Verified professional responses |

#### 3. API Configuration and Access

Data collection used the Python Reddit API Wrapper (PRAW) version 7.6.0, which provides programmatic access to Reddit's official API. Researchers must obtain API credentials through Reddit's developer portal (https://www.reddit.com/prefs/apps). All collection adhered to Reddit's API rate limits (60 requests/minute for OAuth2 clients). A minimum 1-second delay was enforced between requests, with exponential backoff for rate-limit responses (HTTP 429).

#### 4. Collection Parameters

*Table S1.2. Collection parameters and rationale.*

| Parameter | Value | Rationale |
|---|---|---|
| Temporal range | Feb 2019 – Nov 2024 | Pre-pandemic through post-pandemic |
| Sort method | top (all time), hot, new | Balanced representation |
| Minimum score | No threshold | Full narrative diversity |
| Comment depth | Full thread expansion | Complete conversational context |
| Excluded fields | author, user_id, IP data | Privacy preservation |

#### 5. Corpus Characteristics

*Table S1.3. Final corpus characteristics.*

| Characteristic | Value | Details |
|---|---|---|
| Total threads | 5,386 | From 4 subreddits (2019–2024) |
| Total comments | 185,181 | Average thread depth: 34.4 (SD = 47.2) |
| Total words | 2,295,242 | Naturalistic health discourse |
| Topic: Diabetes | 29.0% | Type 1, Type 2, medication management |
| Topic: Obesity/Weight | 24.9% | Diet, exercise, weight loss |
| Topic: Hypertension | 21.9% | Blood pressure, medication adherence |
| Topic: COVID-19 | 16.6% | Symptoms, treatments, prevention |
| Topic: General health | 7.7% | Multi-condition discussions |

#### 6. Text Preprocessing Pipeline

Raw Reddit text underwent systematic cleaning: (1) removal of URLs and hyperlinks; (2) elimination of special characters, emojis, and non-ASCII characters; (3) conversion to lowercase for normalization; (4) removal of Reddit-specific formatting (markdown syntax, user mentions); and (5) elimination of redundant whitespace. Medical abbreviations (e.g., BP, A1C, BG) were preserved to maintain clinical meaning.

#### 7. Stratified Sampling for Annotation

From the master corpus, 2,000 narrative segments were selected using stratified random sampling ensuring proportional representation across health topics and engagement levels. Stratification criteria: (1) subreddit proportional to corpus distribution; (2) balanced sampling across engagement quartiles; (3) temporal distribution matching the full corpus; and (4) minimum segment length of 50 tokens for sufficient narrative structure.

#### 8. Ethical Considerations

The Reddit corpus consists of publicly available posts collected in compliance with Reddit's Terms of Service and API guidelines. No usernames or personally identifying information were collected. All published examples are paraphrased to prevent reverse identification. The research is IRB exempt under 45 CFR 46.104(d)(2). Due to Reddit's content dynamics, exact corpus replication is not possible; researchers should target comparable scale using the same subreddit targets and temporal range.
