## Supplementary File 5 for "VERITAS: A Neuro-Symbolic Approach to Quantifying Epistemic Divergence and Harm Potential in Online Health Narratives"

### Detailed Narrative Case Studies by Quadrant

Corpus quadrant distribution (N = 699 classified narratives): Q1 = 21.5%, Q2 = 14.4%, Q3 = 25.2%, Q4 = 38.9%. Case studies 1, 3, 4, and 5 are drawn from the corpus (lightly condensed for presentation) with their computed NTD/NRS scores; case studies 2 and 6 are illustrative composites with representative scores within the observed ranges (NTD 0.002–0.745; NRS 0.154–0.913).

#### 1. Q3: Aligned but Risky (25.2%) — the safety gap

Epistemically aligned language describing potentially harmful self-management. Structurally invisible to accuracy-based screening.

*Case study 1 (corpus): Deferred response to recurrent chest pain.*

> "It's been an hour of me just resting and the pain is still coming and going. I'm not sure if this is just a concern to bring up next time [with my doctor]."

AAO: Agent = [narrator]; Action = [resting, deferring care]; Outcome = [pain persisting]. Scores: NTD = 0.12 (aligned), NRS = 0.79 (high). No inaccurate claim is made; the risk lies in the invited behavior, treating ongoing chest pain as deferrable. Accuracy-based classifiers approve this content.

*Case study 2 (illustrative): Unsupervised metformin dose escalation.*

> "I've been on metformin 500 mg twice daily for Type 2, but my fasting glucose stayed 160–180 mg/dL. I read that 1000 mg twice daily is common, so I increased it myself. Glucose dropped to 95–110. I'll continue until my appointment in 3 months."

Representative scores: NTD = 0.09, NRS = 0.68. Every terminological element is correct; unsupervised dose doubling without clinical review carries risk (including rare lactic acidosis in predisposed individuals) that no factual check detects.

#### 2. Q4: Divergent & Risky (38.9%)

*Case study 3 (corpus): Self-managed diabetic ketoacidosis.*

> "This happens to me sometimes when I have mild DKA and fix it myself quickly."

Scores: NTD = 0.56 (divergent), NRS = 0.89 (high). Minimizing diabetic ketoacidosis, a medical emergency, as self-manageable diverges from consensus and invites acutely harmful behavior. Both dimensions flag this content; the pipeline's emergency-response detector also fires.

#### 3. Q1: Aligned & Safe (21.5%)

*Case study 4 (corpus): Referral to clinical review.*

> "He needs to see a doctor and have his diabetes control reviewed; if this is the cause it's easily fixed with medications."

Scores: NTD = 0.15, NRS = 0.26. Consensus-aligned advice directing the reader toward clinical care. Both binary and two-dimensional approaches handle this correctly.

#### 4. Q2: Divergent but Safe (14.4%) — the equity gap

*Case study 5 (corpus): Unverified self-observation.*

> "I noticed that I feel this way the most when I've had something sweet and my glucose has jumped."

Scores: NTD = 0.60, NRS = 0.22. An epistemically divergent personal causal attribution with no risky action invited. Accuracy-based screening flags such content as misinformation; the risk dimension assesses it as behaviorally benign, supporting contextualization rather than suppression.

*Case study 6 (illustrative): Traditional remedy as adjunct.*

"Using turmeric paste for joint stiffness instead of NSAIDs. Seems to help and I prefer natural approaches."

Representative scores: NTD = 0.42, NRS = 0.28. Divergent from standard practice yet low harm potential for this self-limited complaint; blanket suppression of such content disproportionately affects culturally traditional practices.

### 5. AAO Graph Structure

Each narrative graph contains Agent nodes (UMLS-mapped), Action nodes (interventions/behaviors), Outcome nodes (results), and Quantifier nodes (clinical measurements). Edge types: PERFORMS (agent→action), RESULTS_IN (action→outcome), TRIGGERS (outcome→action, for emergency-response patterns), HAS_VALUE (quantifier→outcome).
