## Supplementary File 6 for "VERITAS: A Neuro-Symbolic Approach to Quantifying Epistemic Divergence and Harm Potential in Online Health Narratives"

### Weight Optimization and Robustness Validation

#### 1. Weight Sensitivity Analysis

Eleven weight configurations were tested for NTD = $w_F \times D_F + w_C \times D_C + w_E \times D_E$, spanning the primary (theoretically constrained, empirically optimized) configuration, equal weighting, single-pillar emphasis, single-pillar extremes, and leave-one-pillar-out ablations.

*Table S6.1. Configurations tested and NTD–NRS independence (N = 699 classified narratives).*

| Configuration | $D_F$ | $D_C$ | $D_E$ | Pearson r | Spearman ρ | \|r\| < 0.30 |
|---|---|---|---|---|---|---|
| Primary (used) | 0.250 | 0.350 | 0.400 | 0.222 | 0.229 | Yes |
| Equal | 0.333 | 0.333 | 0.334 | 0.209 | 0.212 | Yes |
| Factual Heavy | 0.500 | 0.300 | 0.200 | 0.169 | 0.167 | Yes |
| Causal Heavy | 0.200 | 0.500 | 0.300 | 0.215 | 0.226 | Yes |
| Epistemic Heavy | 0.200 | 0.300 | 0.500 | 0.234 | 0.244 | Yes |
| Extreme $D_F$ | 0.800 | 0.100 | 0.100 | −0.012 | −0.001 | Yes |
| Extreme $D_C$ | 0.100 | 0.800 | 0.100 | 0.204 | 0.215 | Yes |
| Extreme $D_E$ | 0.100 | 0.100 | 0.800 | 0.268 | 0.267 | Yes |
| No $D_F$ | 0.000 | 0.500 | 0.500 | 0.234 | 0.272 | Yes |
| No $D_C$ | 0.500 | 0.000 | 0.500 | 0.131 | 0.125 | Yes |
| No $D_E$ | 0.500 | 0.500 | 0.000 | 0.165 | 0.155 | Yes |

**All 11 configurations maintained |r| < 0.30**, confirming that the practical independence of divergence and harm potential is a property of the constructs rather than of a particular weighting.

#### 2. Quadrant Stability

*Table S6.2. Quadrant distribution (% of 699 classified narratives) under representative configurations. Binary misclassification = Q2 + Q3 (over-flagged safe content plus missed risky content).*

| Configuration | Q1 % | Q2 % | Q3 % | Q4 % | Q2+Q3 % |
|---|---|---|---|---|---|
| Primary (used) | 21.5 | 14.4 | 25.2 | 38.9 | 39.6 |
| Equal | 19.6 | 16.3 | 23.9 | 40.2 | 40.2 |
| Factual Heavy | 16.0 | 19.9 | 20.5 | 43.6 | 40.3 |
| Causal Heavy | 23.9 | 12.0 | 28.2 | 35.9 | 40.2 |
| Extreme $D_C$ | 24.2 | 11.7 | 28.2 | 35.9 | 39.9 |
| Extreme $D_E$ | 14.7 | 21.2 | 17.5 | 46.6 | 38.6 |
| No $D_C$ | 10.2 | 25.8 | 12.4 | 51.6 | 38.2 |

Across configurations, the binary misclassification share (Q2 + Q3) remains within 38.2%–43.6%, demonstrating that the safety gap is not an artifact of the chosen weights.

#### 3. Shapley Value Decomposition

*Table S6.3. Marginal contribution of each pillar to misinformation-class $F_1$ (full-coalition $F_1$ = 0.336; Shapley decomposition over all pillar coalitions).*

| Component | Shapley value | Share | Interpretation |
|---|---|---|---|
| $D_C$ (Causal) | 0.133 | 39.7% | Faulty causal inference is the largest driver of discriminative power |
| $D_E$ (Evidence) | 0.119 | 35.3% | Confidence–evidence miscalibration is a major contributor |
| $D_F$ (Factual) | 0.084 | 25.0% | Terminology-level divergence contributes but is not dominant |

#### 4. Baseline Performance

*Table S6.4. Recall of expert-labeled misinformation (n = 127). Text baselines: stratified 5-fold cross-validation on the 437 labeled posts; VERITAS quadrant mappings: the 435 posts with complete pipeline scores; LLM: claude-sonnet-4-5, temperature 0, majority of three repetitions.*

| System | Recall | Precision | $F_1$ |
|---|---|---|---|
| TF-IDF + LR (default) | 8.7% (11/127) | 61.1% | 0.152 |
| TF-IDF + LR (balanced) | 55.9% (71/127) | 52.2% | 0.540 |
| BioBERT emb + LR (default) | 25.2% (32/127) | 43.8% | 0.320 |
| BioBERT emb + LR (balanced) | 57.5% (73/127) | 42.2% | 0.487 |
| LLM zero-shot (risk instruction) | 30.7% (39/127) | 54.2% | 0.392 |
| LLM zero-shot (divergence instruction) | 32.3% (41/127) | 46.1% | 0.380 |
| VERITAS Risk mapping (Q3+Q4) | 62.2% (79/127) | 29.0% | 0.396 |
| VERITAS Divergence mapping (Q2+Q4) | 52.0% (66/127) | 29.2% | 0.374 |

#### 5. Pipeline Characteristics (v3.4)

*Table S6.5. Narrative graph pipeline statistics.*

| Metric | Value | Detail |
|---|---|---|
| AAO triplets extracted | 4,975 | Mean 7.1 per narrative (704 narratives) |
| Causal inversions detected | 346 segment-level | 137 narratives (19.5%) exhibit reactive causation |
| Orphan nodes recovered | 1,613 | Proximity-based reattachment |
| UMLS concept coverage | 100% | Hybrid string + BioBERT semantic normalization |
| Narratives classified | 699 of 704 | 5 yielded no complete AAO triplet |
